# Expanding access to SARS-CoV-2 IgG and IgM serologic testing using fingerstick whole blood, plasma, and rapid lateral flow assays

**DOI:** 10.1101/2021.04.16.21255130

**Authors:** Mark Anderson, Vera Holzmayer, Ana Vallari, Russell Taylor, James Moy, Gavin Cloherty

## Abstract

Serologic testing for SARS-CoV-2 antibodies can be used to confirm diagnosis, estimate seroprevalence, screen convalescent plasma donors, and assess vaccine efficacy. Several logistical and infrastructure challenges limit access to SARS-CoV-2 serologic testing. Dried blood spot (DBS) samples have been used for serology testing of various diseases in resource-limited settings. We examined the use of DBS samples and capillary blood (fingerstick) plasma collected in Microtainer tubes for SARS-CoV-2 testing with the automated Abbott ARCHITECT™ SARS-CoV-2 IgG (List 6R86) and IgM assays and use of venous whole blood with a prototype PANBIO™ rapid point-of-care lateral flow SARS-CoV-2 IgG assay. The ARCHITECT™ SARS-CoV-2 IgG assay was initially optimized for use with DBS, venous and capillary plasma, and venous whole blood collected from patients with symptoms and PCR-confirmed COVID-19 and negative asymptomatic controls. Assay linearity and reproducibility was confirmed with 3 contrived DBS samples, with sample stability and signal recovery after 14 days at room temperature. ARCHITECT™ SARS-CoV-2 IgG and IgM assay results showed high concordance between fingerstick DBS and venous DBS samples, and between fingerstick DBS and venous whole blood samples (n=61). Discordant results were seen in 3 participants (2 IgG, 1 IgM) who were in the process of seroreversion at the time of sample collection and had results near the assay cutoff. Use of fingerstick plasma collected in Microtainer tubes (n=109) showed 100% concordant results (R^2^=0.997) with matched patient venous plasma on the ARCHITECT™ SARS-CoV-2 IgG assay. High concordance of assay results (92.9% positive, 100% negative) was also observed for the PANBIO™ SARS-CoV-2 IgG assay compared to the ARCHITECT™ SARS-CoV-2 IgG assay run with matched venous plasma (n=61). Fingerstick DBS and plasma samples are easy and inexpensive to collect and, along with the use of rapid point-of-care testing platforms, will expand access to SARS-CoV-2 serology testing, particularly in resource-limited areas.

## INTRODUCTION

Serologic testing for SARS-CoV-2 antibodies can be used to complement PCR-based diagnostic testing [1] and may assist in identifying asymptomatic cases and those with past infection as potential donors for convalescent plasma therapy. The COVID-19 pandemic has presented unique barriers to achieving widespread serologic testing, such as logistical and infrastructure challenges that limit the number of available testing sites, transportation issues that reduce patient access to existing testing sites, and reluctance among patients to seek out testing in clinical settings that may increase risk of exposure to the virus. In addition, current serologic assays utilize venous whole blood or plasma samples that require trained phlebotomists and sophisticated laboratory equipment. Removing these hurdles would improve access to SARS-CoV-2 serologic testing and expand the ability to track the virus and its community spread. Innovative approaches to expanding SARS-CoV-2 serologic testing include the development of fully automated assay systems to increase testing speed and rapid point-of-care testing platforms, as well as the use of new types of samples that can be collected at home or in drive-through settings.

In low- and middle-income countries (LMICs) that lack capacity for venous blood draws or where transporting and storing blood samples is difficult, the use of dried blood spot (DBS) samples has improved access to diagnostic testing for various infectious diseases [2]. DBS samples are collected by applying blood from capillary puncture (fingerstick) directly to an absorbent paper card, which are dried and can be sent through the mail to a centralized lab. The samples are recovered from the card by eluting in buffer. DBS samples are now widely used in low-resource settings for diagnosis and therapeutic monitoring of HIV and HBV [3-5]. Rapid lateral flow assays that utilize capillary blood further simplify serologic testing by combining sample collection and the assay in a single step, reducing the need for additional equipment and training. Capillary plasma can also be collected in Microtainer tubes, which eliminates the need for venipuncture. Previous studies found no difference in IgG serologic assay results when using capillary versus venous plasma samples [6, 7].

In this study, we assessed the feasibility of using capillary DBS samples and capillary plasma as the starting material for the Abbott ARCHITECT™ SARS-CoV-2 IgG assay, approved under Emergency Use Authorization (EUA) to detect IgG antibodies against the SARS-CoV-2 nucleocapsid protein in human serum and plasma, and a prototype ARCHITECT SARS-CoV-2 IgM assay that detects IgM antibodies against the spike protein. We also examined the clinical performance of these assays using DBS samples generated using venous or capillary whole blood compared to venous plasma. Finally, we performed a preliminary evaluation of the Abbott PANBIO™ lateral flow SARS-CoV-2 IgG assay performance compared with the ARCHITECT SARS-CoV-2 IgG assay.

## MATERIALS AND METHODS

### Study design and participants

The DBS study was conducted in two parts. First, contrived DBS samples were used to optimize the ARCHITECT™ SARS-CoV-2 IgG assay parameters. Then a clinical performance study was conducted to compare assay performance with DBS samples generated from capillary blood (fingerstick), DBS samples generated from venous whole blood, and venous plasma as the gold standard. Study participants were patients and employees who presented to Rush University Medical Center with a diagnosis of COVID-19 after a positive SARS-CoV-2 PCR-based diagnostic test or with symptoms suggestive of COVID-19. Samples were also collected from a negative control cohort of participants without COVID-19 symptoms. The study protocol was approved by the Institutional Review Board at Rush University Medical Center (IRB# 20041610-IRB01).

The Microtainer clinical performance study (IRB# 20062506-IRB01) was conducted to compare ARCHITECT™ SARS-CoV-2 IgG assay performance using plasma generated from capillary fingerstick blood and plasma generated from venous blood. Study participants (n=109) were patients and employees of Rush University Medical Center with a diagnosis of COVID-19 after a positive SARS-CoV-2 PCR based diagnostic test, or with symptoms suggestive of COVID-19. Samples were also collected from a negative control cohort of participants without COVID-19 symptoms. After providing informed consent, up to 15 mL of venous blood was collected into vacutainer tubes and fingerstick blood was collected in microtainer tubes from each study participant. Plasma from venous and fingerstick blood samples was isolated. Matched venous and fingerstick plasma samples from each patient were run on the ARCHITECT SARS-CoV-2 IgG assay to obtain comparative index results.

The performance of the PANBIO lateral flow SARS-CoV-2 IgG assay using venous whole blood samples was compared to that of the ARCHITECT SARS-CoV-2 IgG assays run with matched venous plasma. Study samples were collected as described above under IRB# 20041610-IRB01. Testing was performed by pipetting 20 µL of venous whole blood and 20 µL of assay buffer into the appropriate wells of a PANBIO lateral flow cartridge, and results were read after 10 minutes. A Pink/Red control line appears in the C area and a Pink/Red line appears in the G area of the reading window for a Positive result. A Pink/Red control line appears in the C area and No Pink/Red line appears in the G area of the reading window for a Negative result. If there is No Pink/Red control line in the C area, even if a line appears in the G area of the reading window, the result is Invalid. Testing and result reporting was performed blinded, with the test performer having no knowledge of which samples were SARS-CoV-2 IgG positive or negative based on previous ARCHITECT SARS-CoV-2 IgG assays with matched patient venous plasma samples. There were no Invalid test results.

### DBS assay samples

After providing informed consent, up to 12 mL of EDTA venous whole blood was collected from each study participant (n=61) along with fingerstick whole blood. Five separate DBS samples were generated from fingerstick whole blood, and another five DBS samples were generated from venous whole blood. Plasma was separated using 6 mL of venous whole blood and the remaining 6 mL of venous whole blood was stored at -80°C. Deidentified plasma, whole blood, and DBS samples were frozen at -80°C and shipped to Abbott Diagnostics (Abbott Park, IL) for use in serological assays. To evaluate the clinical performance of the IgG and IgM assays using DBS samples, for each participant, assay results were compared for venous whole blood DBS, fingerstick whole blood DBS, and venous plasma as the gold standard.

### DBS serological assays

All samples were run on the Abbott ARCHITECT *i*2000SR instrument using the EUA-approved SARS-CoV-2 IgG (List 6R86) assay and prototype SARS-CoV-2 IgM assay (Abbott Diagnostics, Abbott Park, IL) per the ARCHITECT operations manual and assay package insert instructions, with volume modifications for the DBS samples based on optimization experiments. The qualitative SARS-CoV-2 IgG and IgM assays use chemiluminescent microparticles to detect IgG bound to the SARS-CoV-2 nucleocapsid protein or IgM bound to the SARS-CoV-2 spike protein. Assay results are measured in Relative Light Units (RLU) and reported as an index value of the ratio of specimen to calibrator RLU signal (S/C or S/Co). Index values ≥1.4 S/C indicate a SARS-CoV-2 IgG seropositive result and index values ≥1.0 S/C indicate a SARS-CoV-2 IgM seropositive result. The diagnostic accuracy of the ARCHITECT SARS-CoV-2 IgG assay [8-10] and prototype ARCHITECT SARS-CoV-2 IgM assay [11] have been previously reported.

### DBS assay optimization

For assay volume optimization and stability studies, venous plasma from a commercially available SARS-CoV-2 IgG-positive patient was serially diluted in negative whole blood. DBS samples were generated by pipetting 70 µL of each whole blood dilution to the center of a 12-mm Whatman 903 (GE Healthcare/LabMate) DBS card (5 replicate spots/card) and dried overnight in a sterile hood. DBS samples were then cut/punched out and placed in a 1.5-mL Eppendorf tube to which 300 µL elution buffer (1X PBS pH 7.4, 0.25% Triton X-100) was added. The tubes were placed on a shaker for 1 hour at room temperature, and the eluate was then transferred into a fresh 2mL cryogenic tube. The Whatman paper was squeezed to transfer as much liquid as possible to the new tube. The samples were centrifuged for 2 minutes at 10,000 RCF before placement on the ARCHITECT *i*2000SR and run with the SARS-CoV-2 IgG or IgM assays.

Assay linearity and stability were assessed using contrived DBS samples generated from 3 different commercially available SARS-CoV-2 IgG-positive patients. DBS cards were placed in plastic bags (Minigrip, LabMate) with 1 g silica gel desiccant (Uline) and stored at -20°C, room temperature, or 37°C in an incubator. After 1, 3, 7, 10, and 14 days, the DBS samples were eluted in 300 µl ABT elution buffer and 150 µL of the eluate was run on the ARCHITECT SARS-CoV-2 IgG assay.

Venous plasma and venous whole blood DBS samples from 4 study participants were eluted in PBS with Triton, PBS alone, or SARS-CoV-2 IgG (List 6R86) Assay Diluent to assess reproducibility.

## RESULTS

### ARCHITECT SARS-CoV-2 IgG assay performance with DBS

A commercially available SARS-CoV-2 IgG positive plasma sample (Sample 1) was serially diluted from 1:5 to 1:100 into negative whole blood. DBS were generated by pipetting 70 µl of each dilution onto Whatman 903 paper and left to dry overnight. DBS were eluted and run with modified SARS-CoV-2 IgG assay parameters to assess signal recovery at various sample volume inputs (Figure 1A).

**Figure 1.**
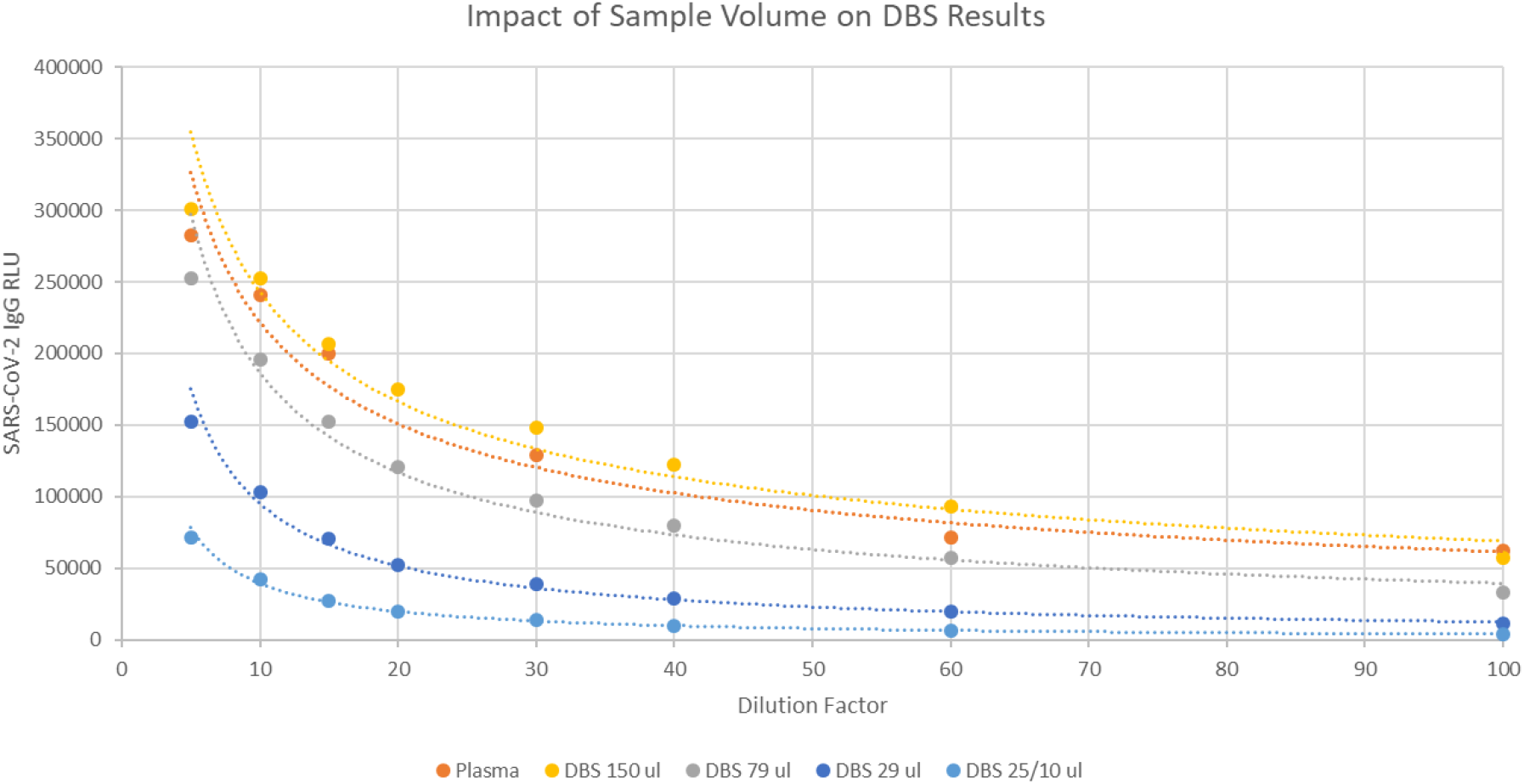
Optimization of ARCHITECT SARS-CoV-2 IgG assay volumes for DBS. (A) Dilution series of DBS samples compared to venous plasma. (B) Linearity of DBS and plasma sample results. (C) Reproducibility of assay results using DBS samples.

DBS assay RLUs were compared to control runs in which the positive IgG sample was diluted into normal human plasma and run using the on-market SARS-CoV-2 IgG assay parameters. A DBS sample volume of 150 µl showed comparable RLU results to the venous plasma sample at each dilution; this volume was used for the remainder of experiments.

Assay linearity and reproducibility were assessed by performing serial dilutions of 3 commercially available SARS-CoV-2 IgG-positive plasma samples in negative whole blood or normal human plasma for DBS and control runs, respectively. DBS testing was performed in triplicate with a 150 µl sample volume and RLUs from the control dilution series were plotted against mean RLU results from the DBS dilutions for Sample 1 (Figure 1B) and Samples 2 and 3 (Supplemental Figure 1A).

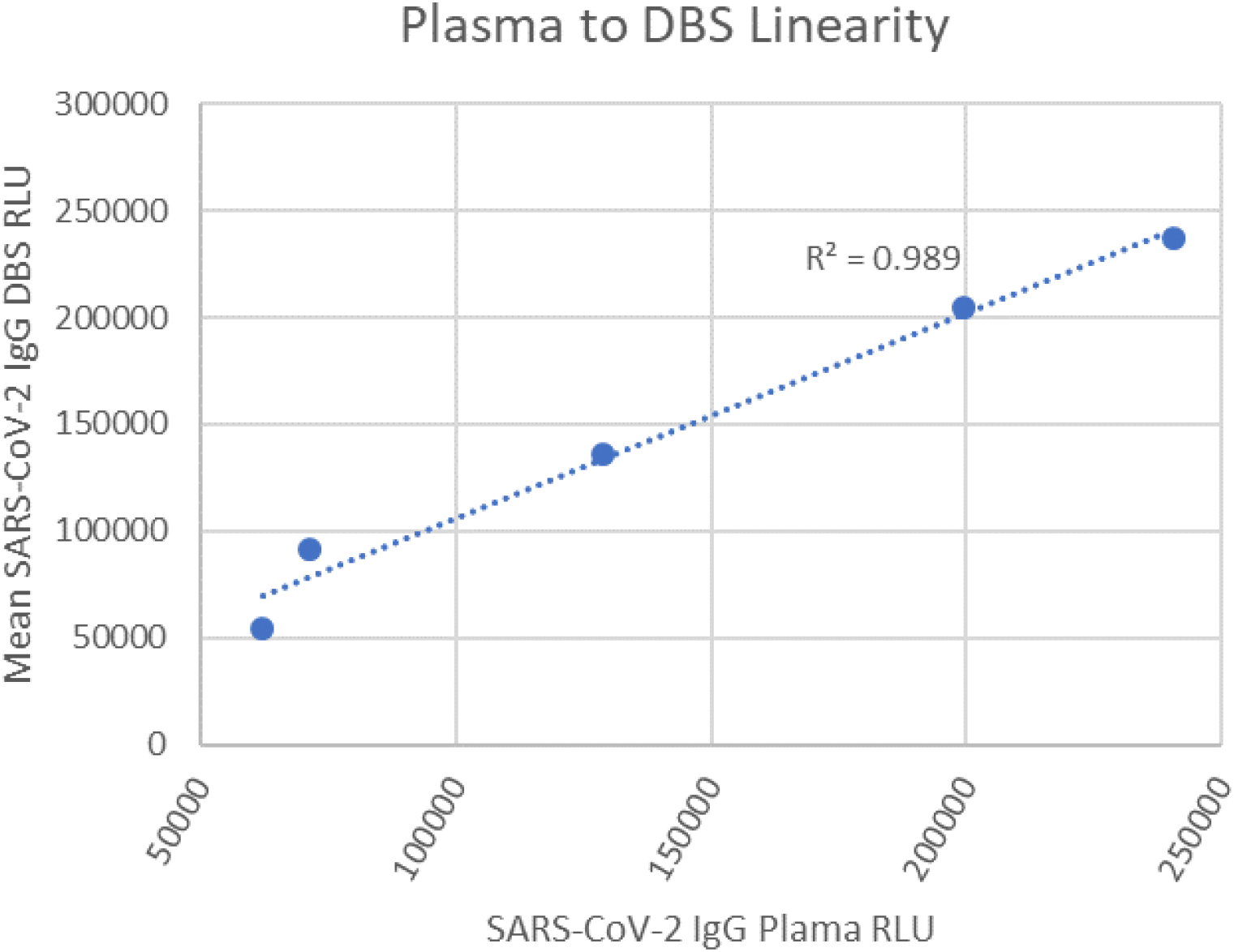

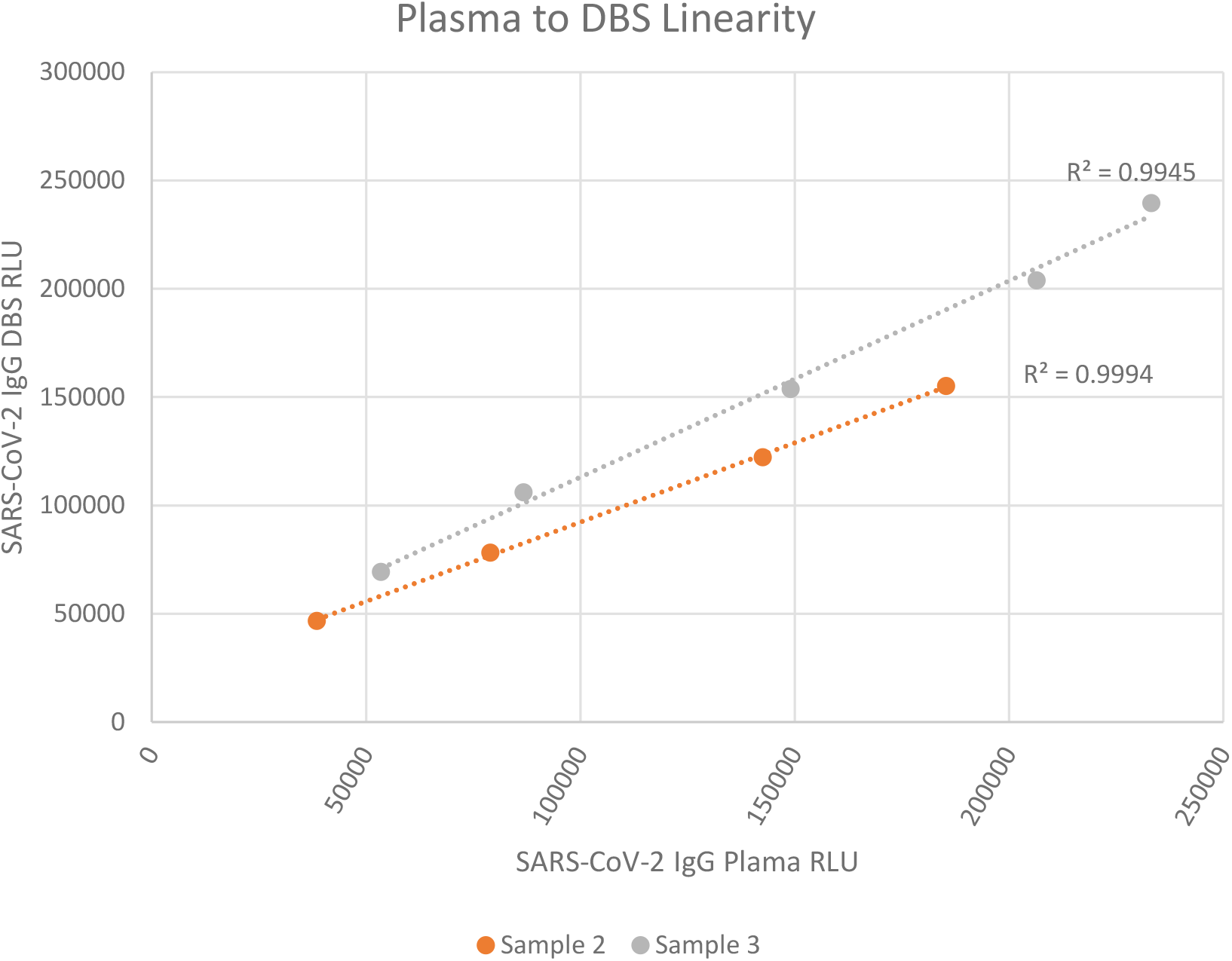

Assay linearity with the 3 samples showed strong correlation (R^2^ = 0.989, 0.995, and 0.999) of the recovered SARS-CoV-2 IgG signal from DBS when compared to plasma controls (Figure 1B and Supplemental Figure 1A). SARS-CoV-2 IgG signal recovery from DBS samples was also shown to be reproducible across all 3 samples (Figure 1C and Supplemental Figure 1B), with %CV values below 6% for all tested conditions except the 1:100 dilution of Sample 1, which had a %CV of 15.33%.

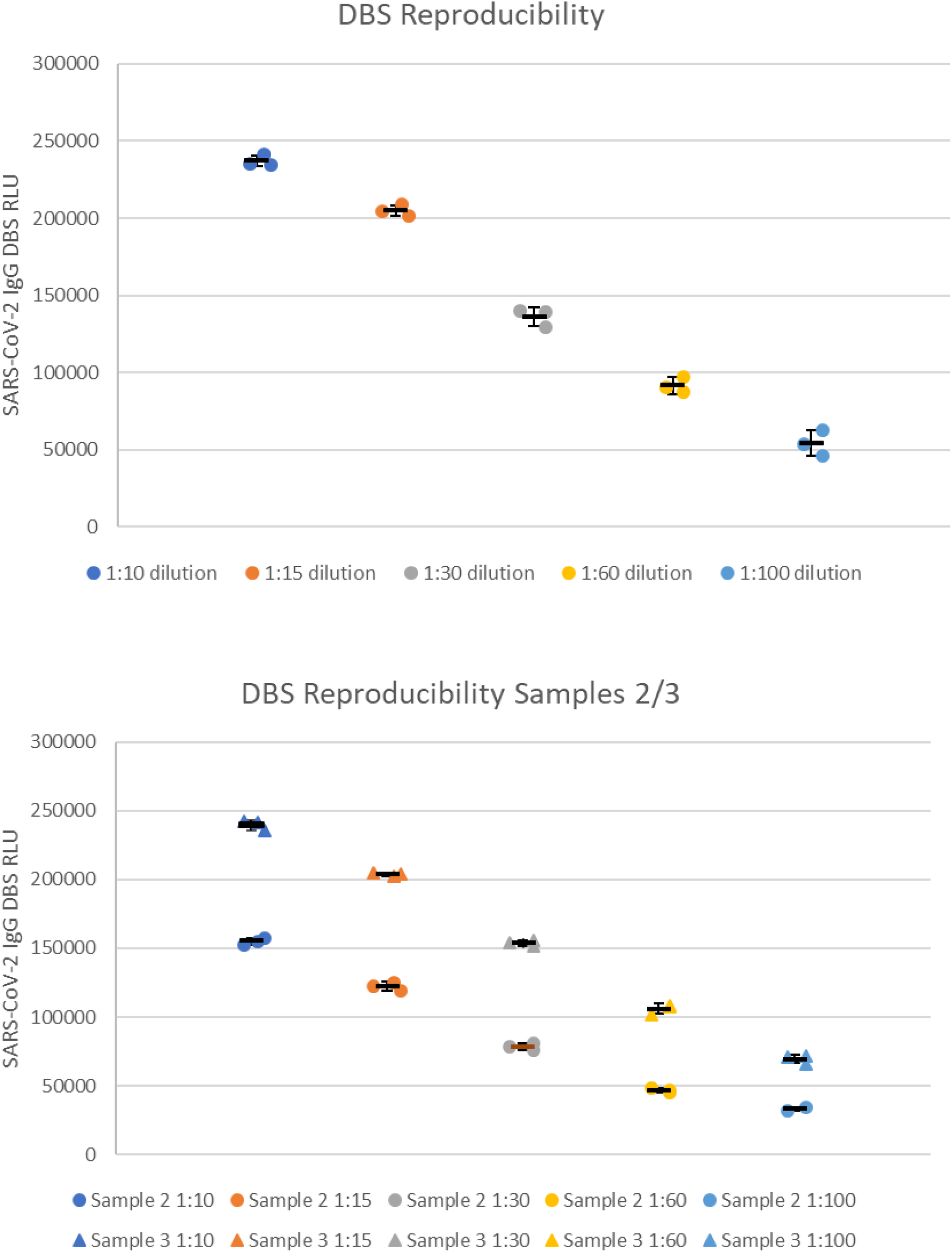

### DBS sample stability

SARS-CoV-2 IgG stability in DBS samples was tested across multiple dilutions for each of the 3 commercially available SARS-CoV-2 IgG-positive samples at room temperature (RT), -20**°**C, and 37**°**C at 1, 3, 7, 10, and 14-day intervals (Table 1: Sample 1, and Supplemental Table 1: Samples 2/3).

**Table 1.**
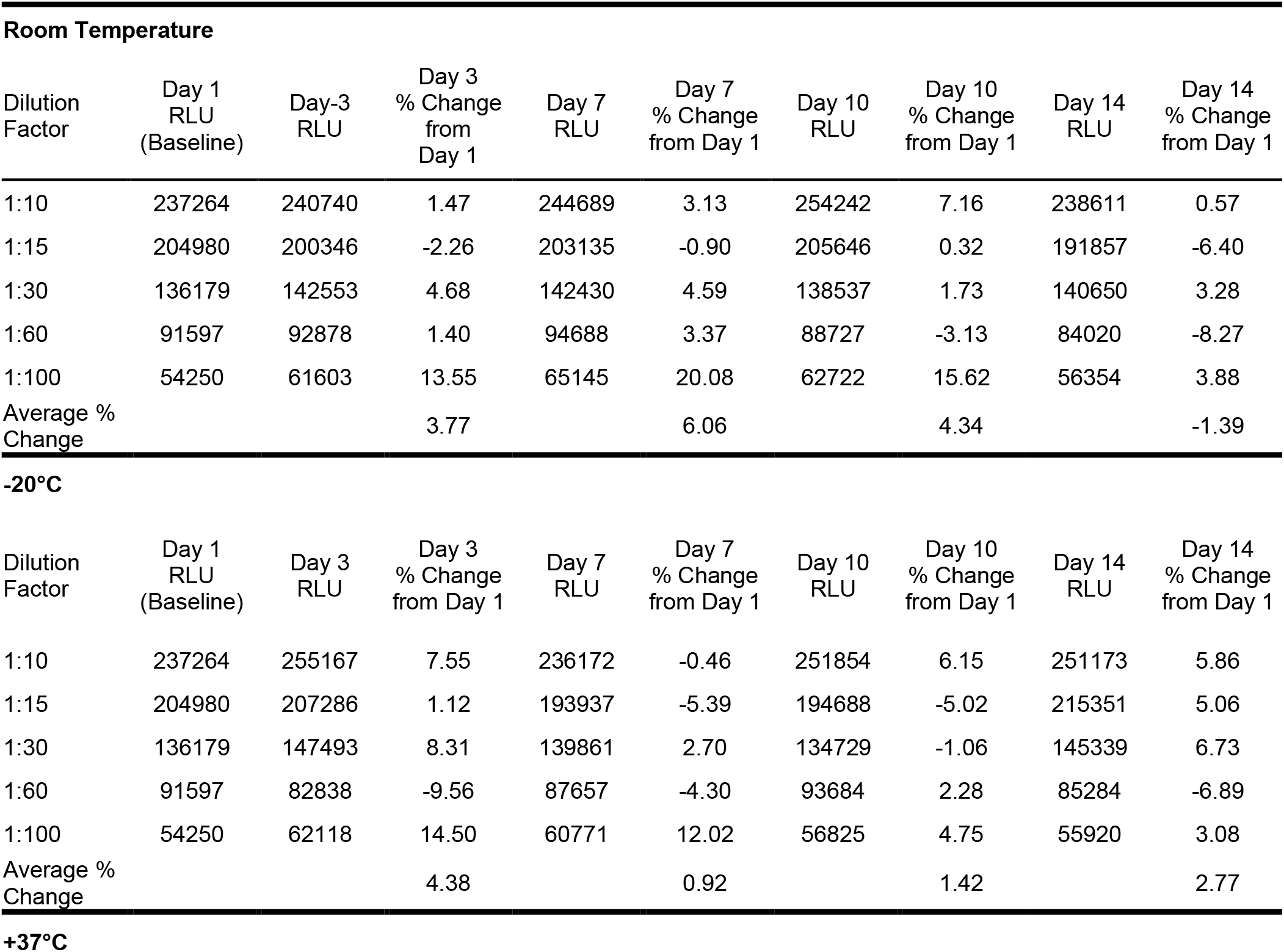
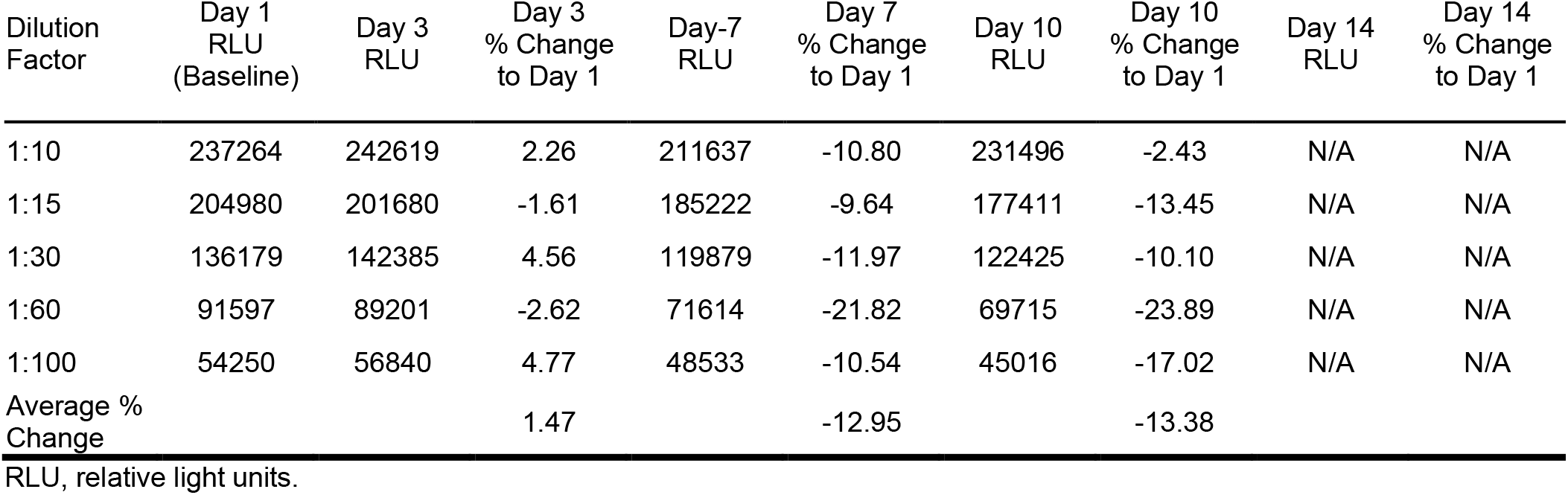
SARS-CoV-2 IgG Results for DBS Sample 1 Stored for Various Times and Temperatures

| **Room Temperature** |  |  |  |  |  |  |  |  |  |
| --- | --- | --- | --- | --- | --- | --- | --- | --- | --- |
| Dilution Factor | Day 1 RLU (Baseline) | Day-3 RLU | Day 3 % Change from Day 1 | Day 7 RLU | Day 7 % Change from Day 1 | Day 10 RLU | Day 10 % Change from Day 1 | Day 14 RLU | Day 14 % Change from Day 1 |
| 1:10 | 237264 | 240740 | 1.47 | 244689 | 3.13 | 254242 | 7.16 | 238611 | 0.57 |
| 1:15 | 204980 | 200346 | -2.26 | 203135 | -0.90 | 205646 | 0.32 | 191857 | -6.40 |
| 1:30 | 136179 | 142553 | 4.68 | 142430 | 4.59 | 138537 | 1.73 | 140650 | 3.28 |
| 1:60 | 91597 | 92878 | 1.40 | 94688 | 3.37 | 88727 | -3.13 | 84020 | -8.27 |
| 1:100 | 54250 | 61603 | 13.55 | 65145 | 20.08 | 62722 | 15.62 | 56354 | 3.88 |
| Average % Change |  |  | 3.77 |  | 6.06 |  | 4.34 |  | -1.39 |
| **-20°C** |  |  |  |  |  |  |  |  |  |
| Dilution Factor | Day 1 RLU (Baseline) | Day 3 RLU | Day 3 % Change from Day 1 | Day 7 RLU | Day 7 % Change from Day 1 | Day 10 RLU | Day 10 % Change from Day 1 | Day 14 RLU | Day 14 % Change from Day 1 |
| 1:10 | 237264 | 255167 | 7.55 | 236172 | -0.46 | 251854 | 6.15 | 251173 | 5.86 |
| 1:15 | 204980 | 207286 | 1.12 | 193937 | -5.39 | 194688 | -5.02 | 215351 | 5.06 |
| 1:30 | 136179 | 147493 | 8.31 | 139861 | 2.70 | 134729 | -1.06 | 145339 | 6.73 |
| 1:60 | 91597 | 82838 | -9.56 | 87657 | -4.30 | 93684 | 2.28 | 85284 | -6.89 |
| 1:100 | 54250 | 62118 | 14.50 | 60771 | 12.02 | 56825 | 4.75 | 55920 | 3.08 |
| Average % Change |  |  | 4.38 |  | 0.92 |  | 1.42 |  | 2.77 |
| **+37°C** |  |  |  |  |  |  |  |  |  |

| Dilution Factor | Day 1 RLU (Baseline) | Day 3 RLU | Day 3 % Change to Day 1 | Day-7 RLU | Day 7 % Change to Day 1 | Day 10 RLU | Day 10 % Change to Day 1 | Day 14 RLU | Day 14 % Change to Day 1 |
| --- | --- | --- | --- | --- | --- | --- | --- | --- | --- |
| 1:10 | 237264 | 242619 | 2.26 | 211637 | -10.80 | 231496 | -2.43 | N/A | N/A |
| 1:15 | 204980 | 201680 | -1.61 | 185222 | -9.64 | 177411 | -13.45 | N/A | N/A |
| 1:30 | 136179 | 142385 | 4.56 | 119879 | -11.97 | 122425 | -10.10 | N/A | N/A |
| 1:60 | 91597 | 89201 | -2.62 | 71614 | -21.82 | 69715 | -23.89 | N/A | N/A |
| 1:100 | 54250 | 56840 | 4.77 | 48533 | -10.54 | 45016 | -17.02 | N/A | N/A |
| Average % Change |  |  | 1.47 |  | -12.95 |  | -13.38 |  |  |

RLU, relative light units.

Day 14 37**°**C data is not available due to an instrument processing error during testing. The average percent change in RLUs across dilution series to the day 1 baselines for each sample showed minimal changes (<±4%) in signal recovery at 14 days at RT and -20**°**C, and moderate signal loss (>±12%) by day 7 at 37**°**C (Table 1 and Supplemental Table 1).

### Clinical performance of the SARS-CoV-2 IgG assay with DBS

A total of 61 participants provided informed consent to have up to 12 ml of venous blood drawn and receive a fingerstick to generate up to 5 DBS samples. An additional 5 (70 µl) DBS samples were generated using a portion of the venous whole blood. Half of the venous whole blood from each study participant was processed to generate a plasma sample and the remaining whole blood was frozen at -80**°**C for future DBS sample generation. Plasma samples from each participant were tested using the ARCHITECT SARS-CoV-2 IgG assay to generate baseline reactivity levels (index value). Fingerstick and venous whole blood DBS samples from each participant were run using the modified SARS-CoV-2 IgG assay (150 µL sample volume). SARS-CoV-2 IgG index value results from fingerstick and venous DBS samples showed good correlation when plotted against index values from plasma (R^2^ = 0.960 and 0.967, respectively; Figure 2A).

**Figure 2.**
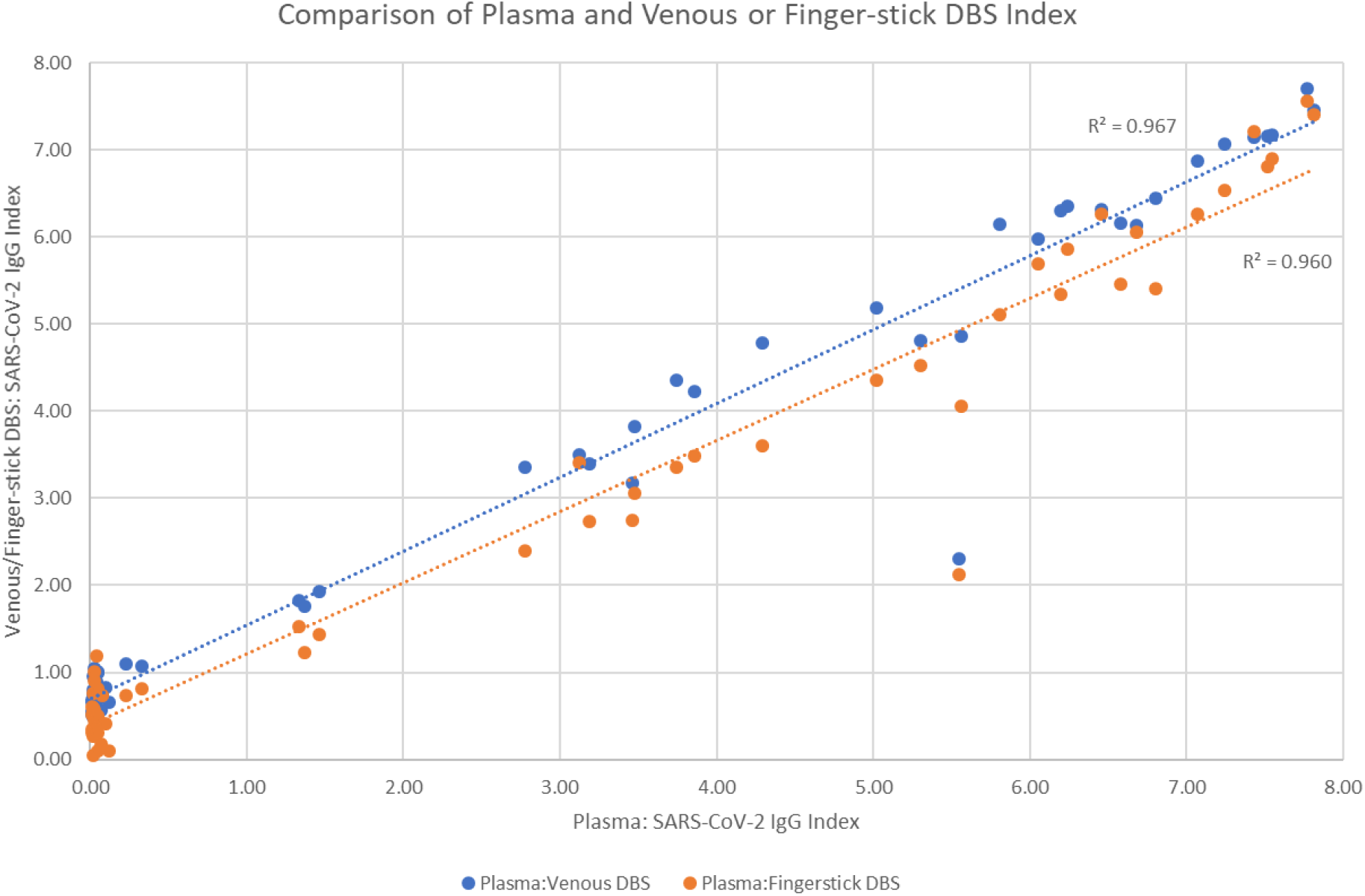
Concordance of ARCHITECT SARS-CoV-2 IgG assay index values for various blood samples. (A) Venous plasma (gold standard) and DBS samples produced from venous or capillary (fingerstick) whole blood. (B) DBS samples produced from venous or capillary (fingerstick) whole blood. DBS samples were eluted in PBS + 0.25% Triton X-100 and 150 µL was used for the IgG assay. (C) Reproducibility of SARS-CoV-2 assay results with venous whole blood DBS. Samples from 2 positive and 2 negative participants were eluted in 300 µL elution buffer and 150 µL was run in triplicate. Standard deviations in the index values were 0.02, 0.04, 0.11, and 0.07 for the samples from these 4 participants.

Further comparison showed that SARS-CoV-2 IgG index values were similar (R^2^ = 0.984) between fingerstick and venous whole blood DBS samples (Figure 2B).

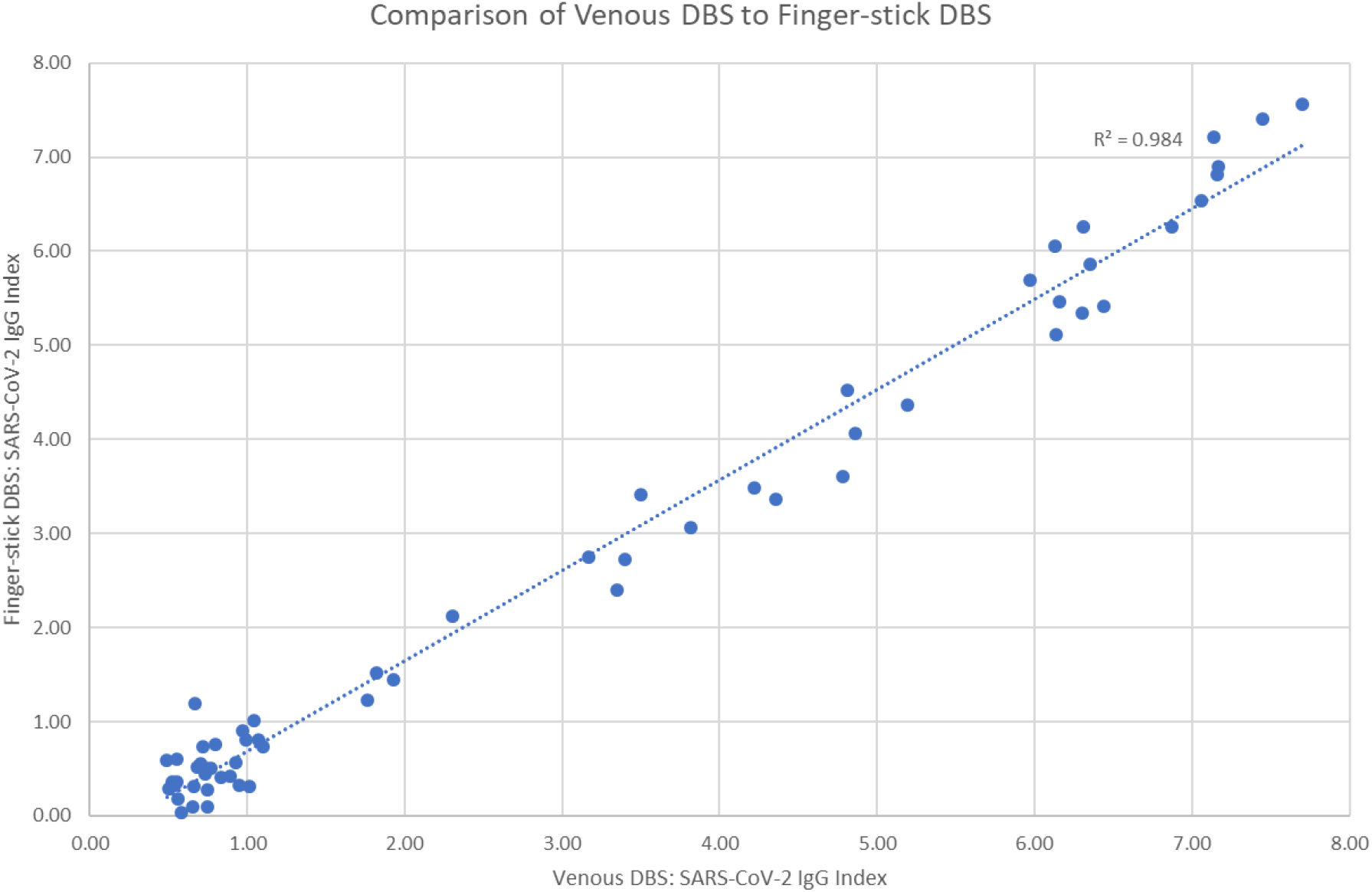

The ARCHITECT SARS-CoV-2 IgG assay utilizes an index cutoff of 1.4 S/C, at or above which a positive result is reported, and below which a negative result is reported. Assay reproducibility was confirmed by testing venous DBS samples from 2 index positive and 2 index negative participants in triplicate (Figure 2C).

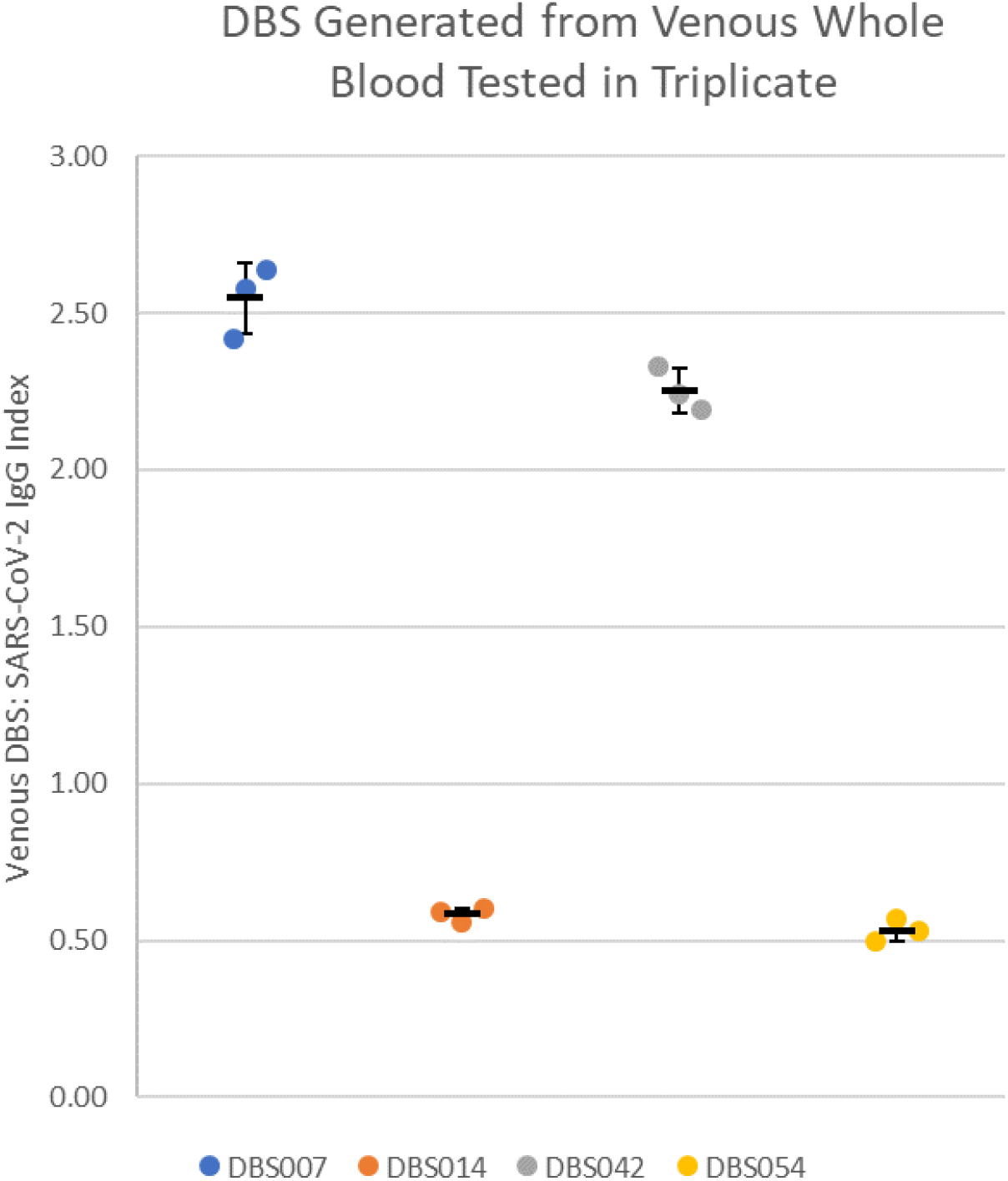

Assay interpretation concordance between reported results for plasma and venous or fingerstick DBS samples was 59/61 (96.7%) and 60/61 (98.4%), respectively (Supplemental Table 2).

### Clinical performance of the SARS-CoV-2 IgM assay with DBS

Patients who are recently infected with SARS-CoV-2 begin producing IgM as early as 5 days after SARS-CoV-2 infection [12]. Serologic testing for SARS-CoV-2 IgM may help confirm SARS-CoV-2 diagnosis in patients who present with symptoms but are negative for PCR-based testing [13]. We examined the performance of the prototype ARCHITECT SARS-CoV-2 IgM assay with DBS samples, using 150 µl of DBS eluate based on our optimization of the SARS-CoV-2 IgG assay. Fingerstick and venous DBS samples from each of the 61 participants were processed and tested with the modified SARS-CoV-2 IgM assay to obtain IgM index values for each patient. Matched patient plasma samples were tested for SARS-CoV-2 IgM using unmodified assay parameters to obtain comparative results. Matched patient index values from plasma plotted against DBS index values (Figure 3A) showed strong correlation for both fingerstick (R^2^ = 0.975) and venous (R^2^ = 0.973) DBS samples.

**Figure 3.**
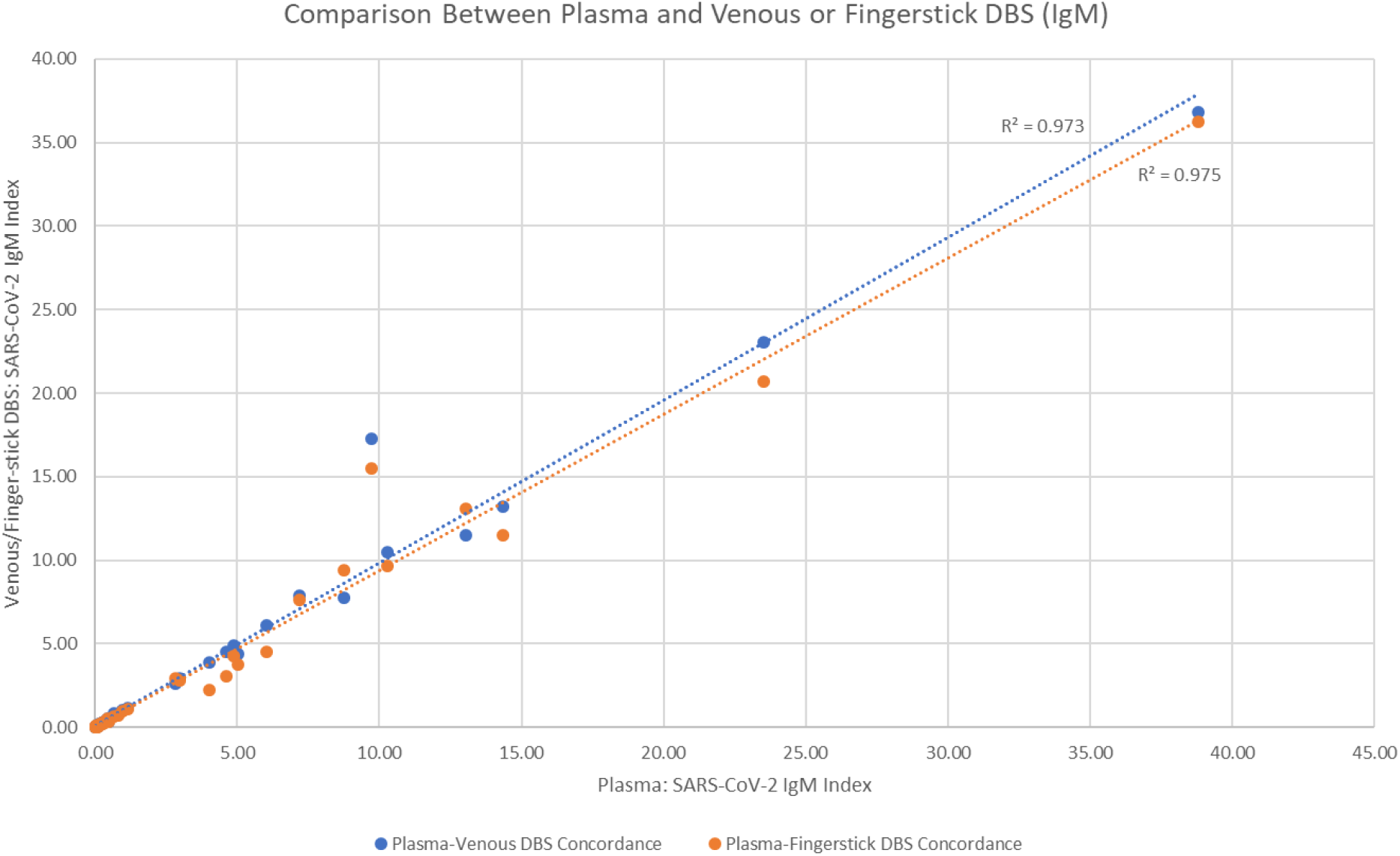
Concordance of ARCHITECT SARS-CoV-2 IgM assay index values for fingerstick and venous whole blood DBS samples and plasma samples. (A) Comparison of plasma index results to venous and fingerstick DBS index results. (B) Equivalency of index values between venous and fingerstick DBS results.

Further comparison showed equivalence (R^2^ = 0.991) between DBS samples generated from fingerstick or venous whole blood (Figure 3B).

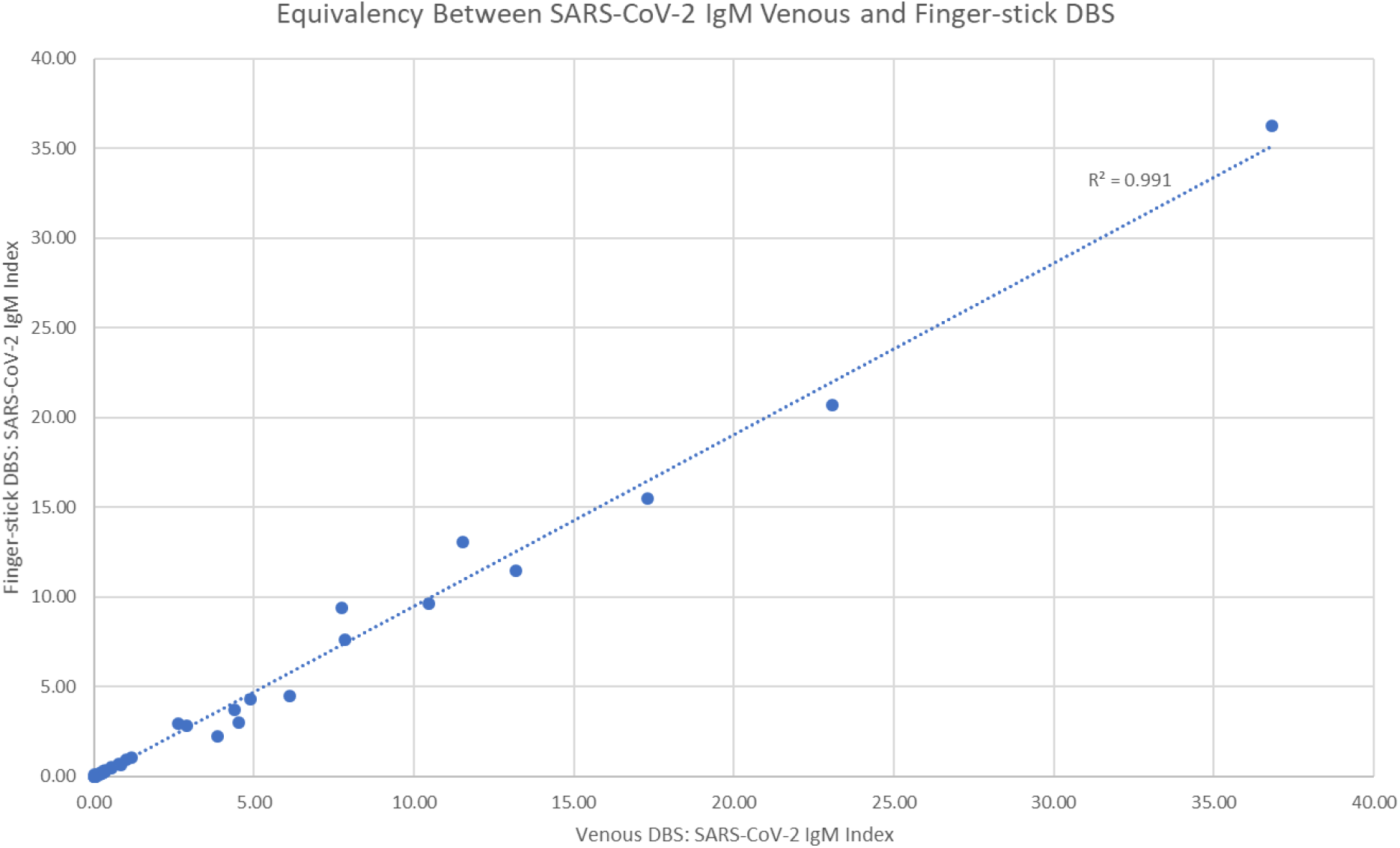

The SARS-CoV-2 IgM assay utilizes an index cutoff of 1.0 S/CO, at or above which a positive result is reported, and below which a negative result is reported. Concordance between reported results for plasma and either venous or fingerstick DBS samples was 60/61 (98.4%) and 61/61 (100%), respectively (Supplemental Table 3).

Similar to the SARS-CoV-2 IgG assay, the single discordant venous DBS sample was from a seroreverting patient whose SARS-CoV-2 IgM plasma index was 0.96, near the assay cutoff.

### Clinical performance of PANBIO SARS-CoV-2 IgG with venous whole blood

Qualitative SARS-CoV-2 IgG reactivity from each of the 61 DBS study participants was determined using retained venous whole blood with the prototype PANBIO lateral flow SARS-CoV-2 assay (Abbott Rapid Diagnostics Jena GmbH). The tester was blinded to which samples were SARS-CoV-2 IgG positive or negative based on previous ARCHITECT studies with matched patient venous plasma. ARCHITECT SARS-CoV-2 IgG positive (index > 1.4) and negative (index < 1.4) concordance with the PANBIO interpretation was 26/28 (92.9%) and 33/33 (100%), respectively (Table 2).

**Table 2.**
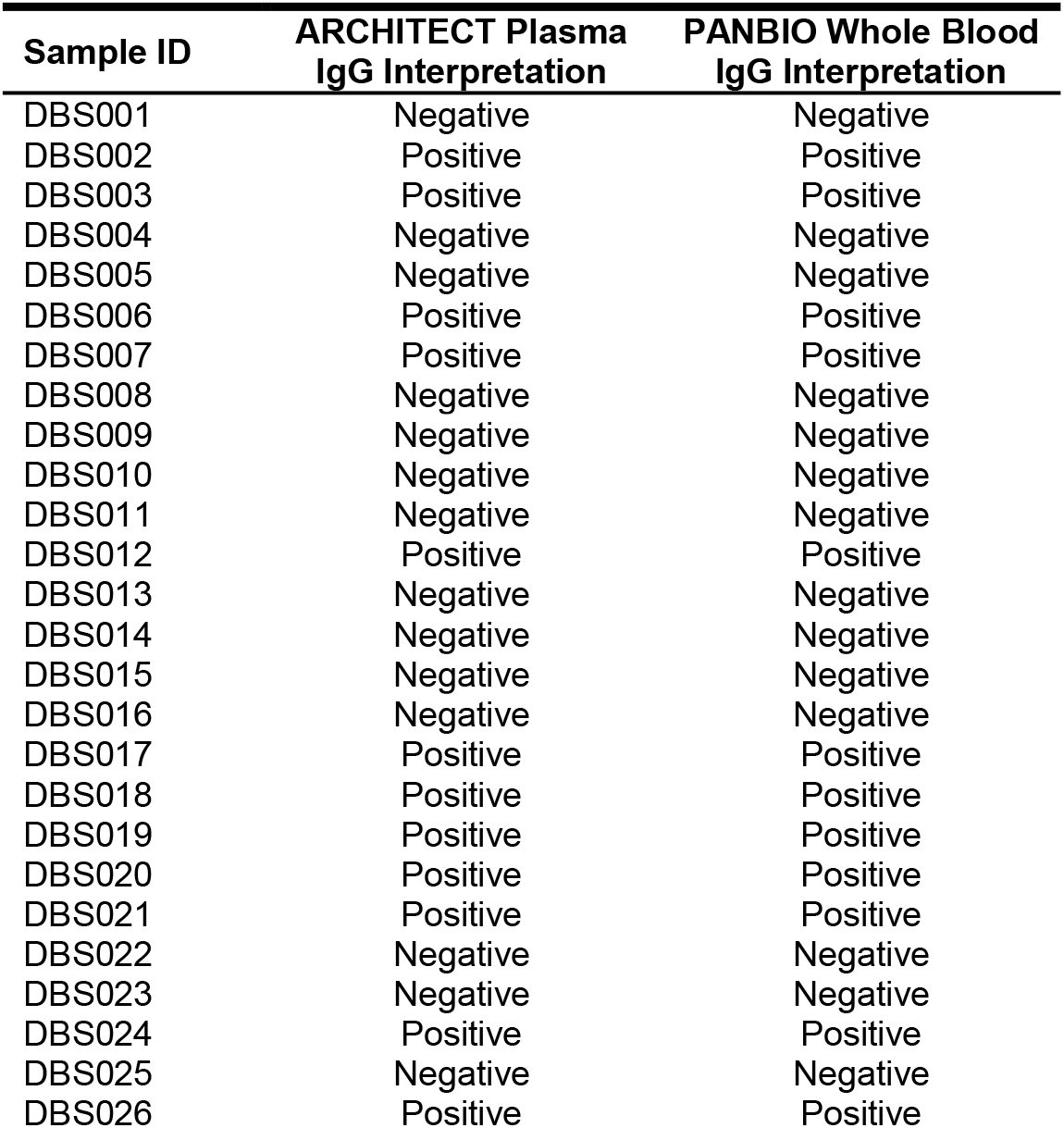
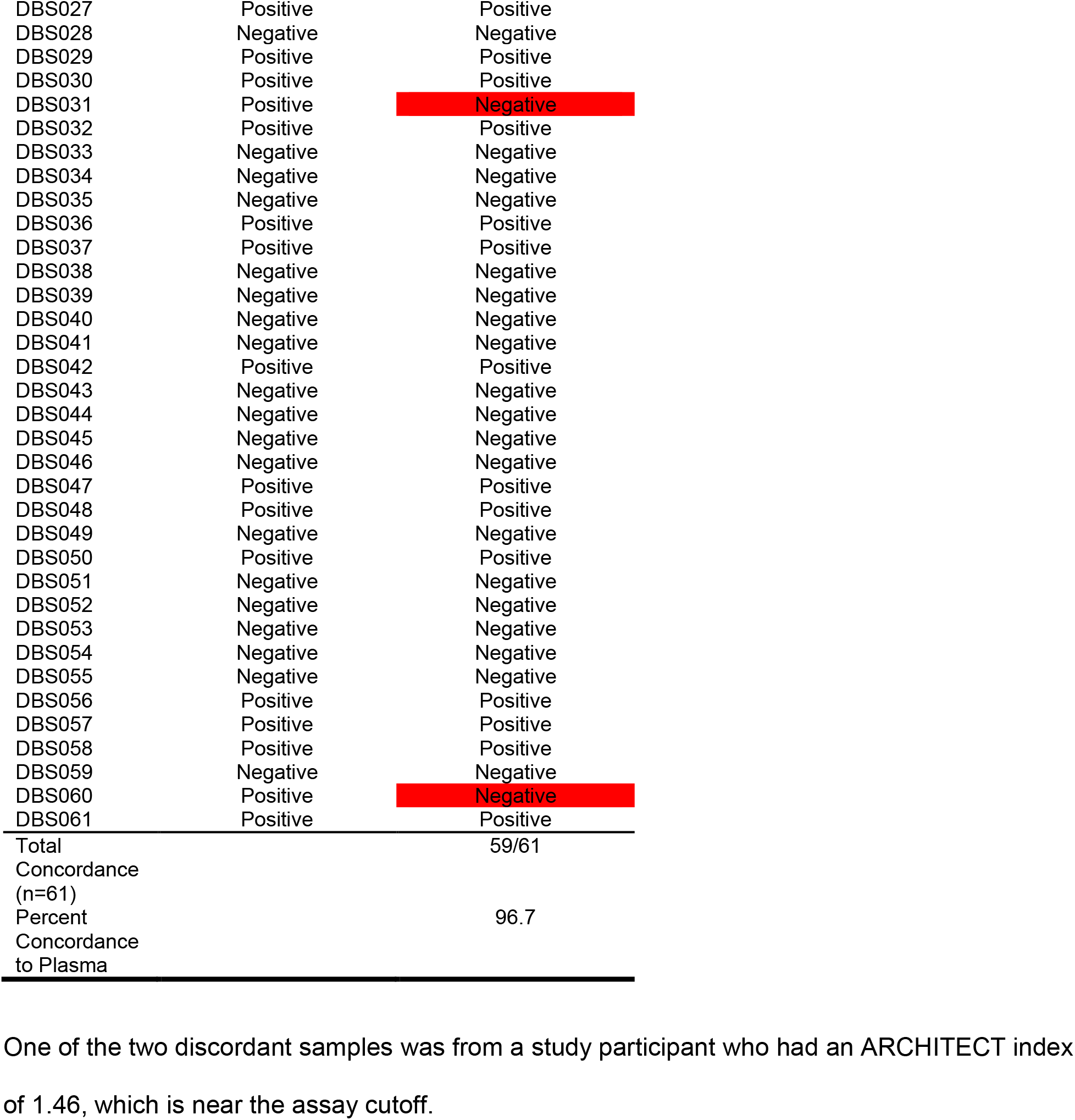
Concordance of Results Interpretation of PANBIO Rapid Point-of-Care SARS-CoV-2 IgG Using Whole Blood and ARCHITECT SARS-CoV-2 IgG Using Venous Plasma

One of the two discordant samples was from a study participant who had an ARCHITECT index of 1.46, which is near the assay cutoff.

### Clinical performance of SARS-CoV-2 IgG using fingerstick plasma

A total of 109 participants provided informed consent to have up to 15 ml of venous blood drawn and receive a fingerstick to generate matched venous and fingerstick plasma samples collected in Vacutainer and Microtainer tubes, respectively. One participant was excluded due to insufficient fingerstick plasma collected. Matched samples were run using the ARCHITECT SARS-CoV-2 IgG assay and the resulting index values were plotted against each other (Figure 4).

**Figure 4.**
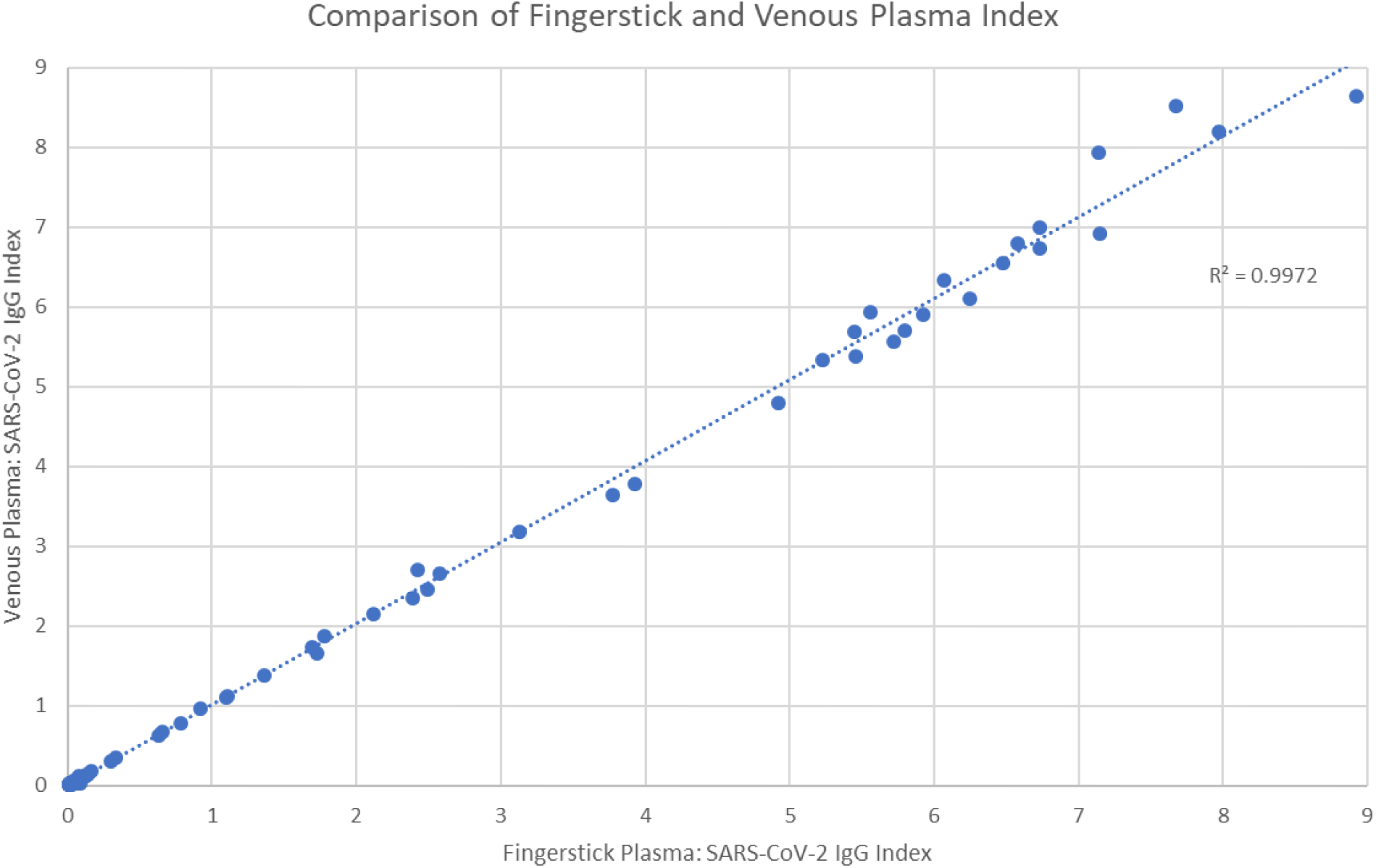
Concordance of ARCHITECT SARS-CoV-2 IgG assay index values for matched fingerstick and venous plasma samples. Fingerstick whole blood was collected in Microtainers and centrifuged to obtain fingerstick plasma. Matched venous plasma was collected during the same visit and both plasma samples were tested for SARS-CoV-2 IgG. Index values from fingerstick and venous plasma were plotted to show a linear relationship.

Correlation between SARS-CoV-2 IgG index values from fingerstick and venous plasma was high (R^2^ = 0.997), and index interpretation concordance was 100% (108/108), demonstrating equivalency between plasma obtained from fingerstick or venous sources.

## DISCUSSION

We investigated different methods to expand and improve access to SARS-CoV-2 antibody testing using sample types that require less processing and handling (DBS, Microtainer plasma, and whole blood) than current methods that use venous plasma and require trained phlebotomists. Our results confirm the feasibility of using DBS as starting material for SARS-CoV-2 IgG and IgM detection on the automated ARCHITECT platform. We found good concordance between the DBS and venous plasma index values on the ARCHITECT SARS-CoV-2 IgG and IgM assays. SARS-CoV-2 IgG DBS samples had higher background in known negative samples, but the results were below the index threshold. Future experiments with different elution conditions will be conducted to further optimize background reduction. Importantly, DBS samples generated from whole venous blood and from fingerstick blood produced concordant assay results, and DBS samples were stable for 2 weeks at room temperature and 1 week at 37°C. Notably, two participants had negative IgG assay results for venous plasma but positive results with DBS samples. The 2 participants with discordant plasma and DBS IgG results and one participant with discordant IgM results had previous positive PCR and IgG results and were found to be in the process of seroreversion at the time the sample used in this study was collected. Thus, these are not false positives with the DBS sample type and may in fact illustrate an increased sensitivity of the DBS assay relative to the plasma assay with the assays’ index value cutoffs. These findings have important implications for expanding access to SARS-CoV-2 serologic testing to areas with low capacity for venous blood draws or lack of refrigerated sample storage.

Our findings are consistent with previous reports on the utility of DBS samples for SARS-CoV-2 serologic testing. Thevis et al. recently described the use of 20 µL capillary blood DBS, collected by fingerstick from 28 participants, 16 of whom had a positive diagnosis by PCR-based testing, for assessment of IgG and IgM antibodies in a lateral flow assay and by ELISA [14]. Comparable results were obtained with DBS samples in the lateral flow and ELISA systems and when compared with test results using conventional serum or plasma samples. A greater number of mismatched results were seen with the IgM assays compared with IgG assays. McDade and colleagues demonstrated the use of DBS samples for SARS-CoV-2 IgG testing of 232 front-line essential workers and their household members to assess seroprevalence in this high-risk population [15].

DBS have been used in PCR and serologic testing for both diagnosis and monitoring of other viral infections, such as HIV, HBV, and HCV, thereby extending testing to rural and remote at-risk populations worldwide [2]. A previous study confirmed the utility of DBS samples with the ARCHITECT HIV, HCV, and HBV serologic assays, reporting specificity of 100% and sensitivities ranging from 97% to 100% [16]. Karp et al. also recently reported the development of a kit for at-home collection of fingerstick DBS for SARS-CoV-2 IgG PCR-based diagnostic testing to a central lab [17], with 100% specificity and sensitivity. The willingness to collect and prepare DBS samples at home and ship them to a central lab for SARS-CoV-2 testing and immune status, was recently confirmed in a survey study of 153 US adults [18]. The ability to use DBS samples collected at home will help overcome current logistical barriers and expand access to SARS-CoV-2 serologic testing in the US and worldwide.

This study also demonstrated equivalent SARS-CoV-2 IgG assay results using plasma collected by fingerstick or by venipuncture. Fingerstick plasma has been used to detect IgG for the confirmation of celiac disease diagnosis [19] and to detect both IgG and IgM after suspected measles or Rubella infection [7, 20]. Collection of fingerstick plasma in Microtainer tubes is quick and easy and does not require a trained phlebotomist, making it a feasible alternative to venipuncture for use in SARS-CoV-2 curbside testing or drive-through testing sites.

Finally, we have shown good concordance between ARCHITECT SARS-CoV-2 IgG assay run with plasma and the PANBIO rapid point-of-care lateral flow SARS-CoV-2 IgG assay using whole blood. The ability to rapidly, accurately, and affordably determine seroprevalence in a population will be an important tool in the growing arsenal of SARS-CoV-2 diagnostic testing, particularly in resource-restricted areas of the world. The simplicity of performing the lateral flow assay with whole blood also eliminates the need for centrifugation and plasma separation steps, further reducing cost and complexity of obtaining a test result. Future studies will verify the accuracy of the PANBIO SARS-CoV-2 IgG assay using capillary whole blood.

Expanding access to SARS-CoV-2 antibody testing will likely require a combination of different testing methods. We have shown that expanding testing capabilities using DBS, Microtainers, and rapid point-of-care tests is feasible and that results delivered with these methods are comparable to current testing approaches.

## Data Availability

The Data that supports the findings of this study are available from Abbott Laboratories but restrictions apply to the availability of these data. Data are available from the authors upon reasonable request and with the permission of Abbott Laboratories.

## AUTHOR CONTRIBUTIONS

MA designed and performed DBS experiments, analyzed the data, and wrote the manuscript. VH performed feasibility and stability DBS experiments and analyzed the data. AV performed the PANBIO testing, RT created modified ARCHITECT assay files, JM performed the specimen collection and analyzed the data. GC directed the research and analyzed the data. All authors reviewed the manuscript.

---

**Supplemental Table 1.** SARS-CoV-2 IgG Results for DBS Samples 2 and 3 Stored for Various Times and Temperatures

**Room Temperature**
| Dilution Factor | Day 1 RLU (Baseline) | Day 3 RLU | Day 3 % Change to Day 1 | Day 7 RLU | Day 7 % Change to Day 1 | Day 10 RLU | Day 10 % Change to Day 1 | Day 14 RLU | Day 14 % Change to Day 1 |
| --- | --- | --- | --- | --- | --- | --- | --- | --- | --- |
| 1:10 | 155198 | 160009 | 3.10 | 156184 | 0.64 | 158252 | 1.97 | 158153 | 1.90 |
| 1:15 | 122269 | 127802 | 4.52 | 123272 | 0.82 | 121656 | -0.50 | 123227 | 0.78 |
| 1:30 | 78267 | 79029 | 0.97 | 81569 | 4.22 | 82677 | 5.63 | 77584 | -0.87 |
| 1:60 | 46612 | 53484 | 14.74 | 51242 | 9.93 | 46693 | 0.17 | 43701 | -6.25 |
| 1:100 | 33001 | 40784 | 23.59 | 36533 | 10.70 | 40883 | 23.89 | 31116 | -5.71 |
| Average % Change |  |  | 9.39 |  | 5.26 |  | 6.23 |  | -2.03 |

**-20°C**
| Dilution Factor | Day 1 RLU (Baseline) | Day 3 RLU | Day 3 % Change to Day 1 | Day 7 RLU | Day 7 % Change to Day 1 | Day 10 RLU | Day 10 % Change to Day 1 | Day 14 RLU | Day 14 % Change to Day 1 |
| --- | --- | --- | --- | --- | --- | --- | --- | --- | --- |
| 1:10 | 155198 | 161767 | 4.23 | 158668 | 2.24 | 170374 | 9.78 | 153203 | -1.29 |
| 1:15 | 122269 | 126733 | 3.65 | 135344 | 10.69 | 127981 | 4.67 | 124583 | 1.89 |
| 1:30 | 78267 | 76782 | -1.90 | 77920 | -0.44 | 70242 | -10.25 | 78305 | 0.05 |
| 1:60 | 46612 | 45825 | -1.69 | 49319 | 5.81 | 41205 | -11.60 | 44063 | -5.47 |
| 1:100 | 33001 | 34328 | 4.02 | 34687 | 5.11 | 28777 | -12.80 | 30973 | -6.14 |
| Average % Change |  |  | 1.66 |  | 4.68 |  | -4.04 |  | -2.19 |

**+37°C**
| Dilution Factor | Day 1 RLU (Baseline) | Day 3 RLU | Day 3 % Change to Day 1 | Day 7 RLU | Day 7 % Change to Day 1 | Day 10 RLU | Day 10 % Change to Day 1 | Day 14 RLU | Day 14 % Change to Day 1 |
| --- | --- | --- | --- | --- | --- | --- | --- | --- | --- |
| 1:10 | 155198 | 153701 | -0.96 | 140870 | -9.23 | 137566 | -11.36 | N/A | N/A |
| 1:15 | 122269 | 110491 | -9.63 | 113106 | -7.49 | 107433 | -12.13 | N/A | N/A |
| 1:30 | 78267 | 78856 | 0.75 | 68268 | -12.78 | 63980 | -18.25 | N/A | N/A |
| 1:60 | 46612 | 49358 | 5.89 | 34660 | -25.64 | 42420 | -8.99 | N/A | N/A |
| 1:100 | 33001 | 38304 | 16.07 | 22713 | -31.17 | 28145 | -14.71 | N/A | N/A |
| Average % Change |  |  | 2.42 |  | -17.26 |  | -13.09 |  |  |

**Sample 3** **Room Temperature**
| Dilution Factor | Day 1 RLU (Baseline) | Day 3 RLU | Day 3 % Change to Day 1 | Day 7 RLU | Day 7 % Change to Day 1 | Day 10 RLU | Day 10 % Change to Day 1 | Day 14 RLU | Day 14 % Change to Day 1 |
| --- | --- | --- | --- | --- | --- | --- | --- | --- | --- |
| 1:10 | 239548 | 234235 | -2.22 | 237921 | -0.68 | 229585 | -4.16 | 224913 | -6.11 |
| 1:15 | 203923 | 212768 | 4.34 | 201590 | -1.14 | 204345 | 0.21 | 203910 | -0.01 |
| 1:30 | 153708 | 152140 | -1.02 | 140951 | -8.30 | 153486 | -0.14 | 145744 | -5.18 |
| 1:60 | 105968 | 103054 | -2.75 | 102193 | -3.56 | 103781 | -2.06 | 99598 | -6.01 |
| 1:100 | 69365 | 75307 | 8.57 | 69062 | -0.44 | 74985 | 8.10 | 70411 | 1.51 |
| Average % Change |  |  | 1.38 |  | -2.82 |  | 0.39 |  | -3.16 |

**- 20°C**
| Dilution Factor | Day 1 RLU (Baseline) | Day 3 RLU | Day 3 % Change to Day 1 | Day 7 RLU | Day 7 % Change to Day 1 | Day 10 RLU | Day 10 % Change to Day 1 | Day 14 RLU | Day 14 % Change to Day 1 |
| --- | --- | --- | --- | --- | --- | --- | --- | --- | --- |
| 1:10 | 239548 | 232119 | -3.10 | 225612 | -5.82 | 236359 | -1.33 | 228690 | -4.53 |
| 1:15 | 203923 | 199371 | -2.23 | 201422 | -1.23 | 210514 | 3.23 | 214023 | 4.95 |
| 1:30 | 153708 | 153231 | -0.31 | 152936 | -0.50 | 150203 | -2.28 | 154392 | 0.44 |
| 1:60 | 105968 | 104353 | -1.52 | 98284 | -7.25 | 108156 | 2.07 | 101594 | -4.13 |
| 1:100 | 69365 | 72157 | 4.03 | 69244 | -0.17 | 64773 | -6.62 | 64843 | -6.52 |
| Average % Change |  |  | -0.63 |  | -2.99 |  | -0.99 |  | -1.96 |

**+37°C**
| Dilution Factor | Day 1 RLU (Baseline) | Day 3 RLU | Day 3 % Change to Day 1 | Day 7 RLU | Day 7 % Change to Day 1 | Day 10 RLU | Day 10 % Change to Day 1 | Day 14 RLU | Day 14 % Change to Day 1 |
| --- | --- | --- | --- | --- | --- | --- | --- | --- | --- |
| 1:10 | 239548 | 226436 | -5.47 | 221869 | -7.38 | 206328 | -13.87 | N/A | N/A |
| 1:15 | 203923 | 190962 | -6.36 | 190767 | -6.45 | 181641 | -10.93 | N/A | N/A |
| 1:30 | 153708 | 146283 | -4.83 | 128277 | -16.55 | 128851 | -16.17 | N/A | N/A |
| 1:60 | 105968 | 100635 | -5.03 | 89604 | -15.44 | 79097 | -25.36 | N/A | N/A |
| 1:100 | 69365 | 69728 | 0.52 | 61991 | -10.63 | 58208 | -16.08 | N/A | N/A |
| Average % Change |  |  | -4.23 |  | -11.29 |  | -16.48 |  |  |

RLU, relative light units.

**Supplemental Table 2.**
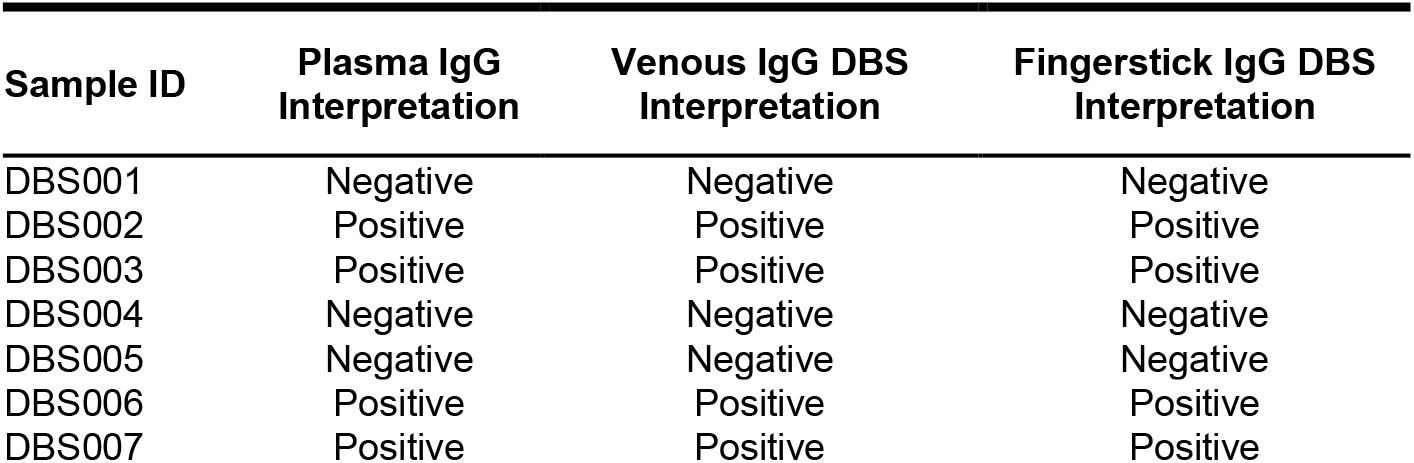
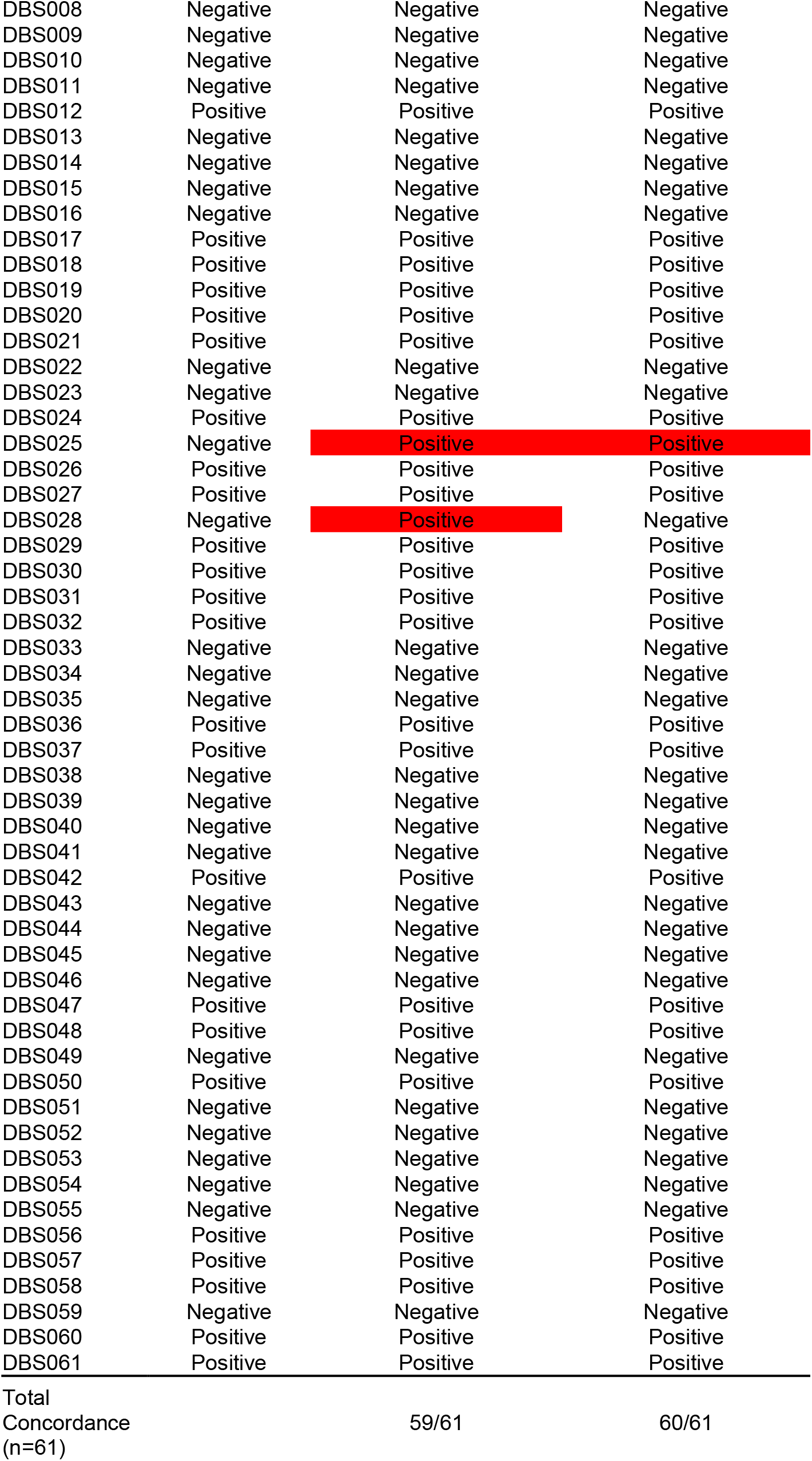
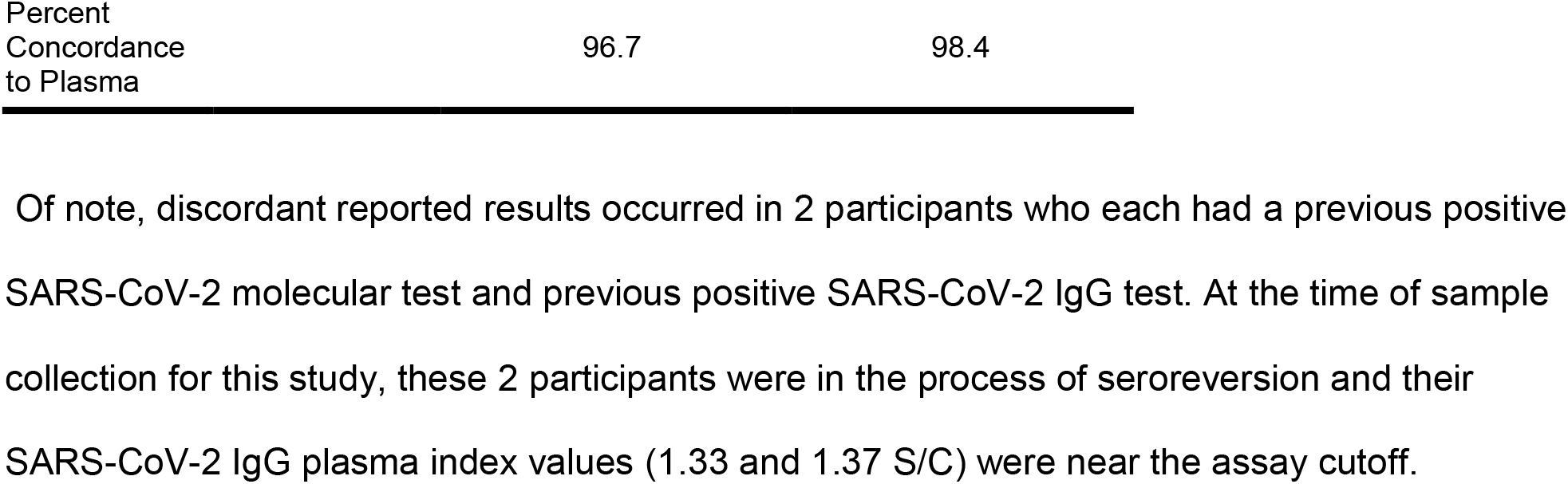
Index Value Concordance Between Plasma, Venous DBS, and Fingerstick DBS on the ARCHITECT SARS-CoV-2 IgG Assay

**Supplemental Table 3.** Index Value Concordance Between Plasma, Venous DBS, and Fingerstick DBS on the ARCHITECT SARS-CoV-2 IgM Assay

| Sample ID | Plasma IgM Interpretation | Venous IgM DBS Interpretation | Fingerstick IgM DBS Interpretation |
| --- | --- | --- | --- |
| DBS001 | Negative | Negative | Negative |
| DBS002 | Positive | Positive | Positive |
| DBS003 | Negative | Negative | Negative |
| DBS004 | Negative | Negative | Negative |
| DBS005 | Negative | Negative | Negative |
| DBS006 | Positive | Positive | Positive |
| DBS007 | Negative | Negative | Negative |
| DBS008 | Negative | Negative | Negative |
| DBS009 | Negative | Negative | Negative |
| DBS010 | Negative | Negative | Negative |
| DBS011 | Negative | Negative | Negative |
| DBS012 | Negative | Negative | Negative |
| DBS013 | Negative | Negative | Negative |
| DBS014 | Negative | Negative | Negative |
| DBS015 | Negative | Negative | Negative |
| DBS016 | Negative | Negative | Negative |
| DBS017 | Negative | Negative | Negative |
| DBS018 | Positive | Positive | Positive |
| DBS019 | Positive | Positive | Positive |
| DBS020 | Negative | Negative | Negative |
| DBS021 | Positive | Positive | Positive |
| DBS022 | Negative | Negative | Negative |
| DBS023 | Negative | Negative | Negative |
| DBS024 | Negative | Negative | Negative |
| DBS025 | Negative | Positive | Negative |
| DBS026 | Positive | Positive | Positive |
| DBS027 | Negative | Negative | Negative |
| DBS028 | Positive | Positive | Positive |
| DBS029 | Negative | Negative | Negative |
| DBS030 | Positive | Positive | Positive |
| DBS031 | Negative | Negative | Negative |
| DBS032 | Positive | Positive | Positive |
| DBS033 | Negative | Negative | Negative |
| DBS034 | Negative | Negative | Negative |
| DBS035 | Negative | Negative | Negative |
| DBS036 | Positive | Positive | Positive |
| DBS037 | Positive | Positive | Positive |
| DBS038 | Negative | Negative | Negative |
| DBS039 | Negative | Negative | Negative |
| DBS040 | Negative | Negative | Negative |
| DBS041 | Negative | Negative | Negative |
| DBS042 | Negative | Negative | Negative |
| DBS043 | Negative | Negative | Negative |
| DBS044 | Negative | Negative | Negative |
| DBS045 | Negative | Negative | Negative |
| DBS046 | Negative | Negative | Negative |
| DBS047 | Positive | Positive | Positive |
| DBS048 | Negative | Negative | Negative |
| DBS049 | Negative | Negative | Negative |
| DBS050 | Positive | Positive | Positive |
| DBS051 | Negative | Negative | Negative |
| DBS052 | Negative | Negative | Negative |
| DBS053 | Negative | Negative | Negative |
| DBS054 | Negative | Negative | Negative |
| DBS055 | Negative | Negative | Negative |
| DBS056 | Positive | Positive | Positive |
| DBS057 | Negative | Negative | Negative |
| DBS058 | Positive | Positive | Positive |
| DBS059 | Negative | Negative | Negative |
| DBS060 | Negative | Negative | Negative |
| DBS061 | Positive | Positive | Positive |
| Total |  |  |  |
| Concordance<br>(n=61) |  | 60/61 | 61/61 |
| Percent<br>Concordance<br>to Plasma |  | 98.4 | 100.0 |

